# Strain coexistence and competition for pathogens with asymmetric cross-immunity and waning immunity

**DOI:** 10.64898/2026.08.03.26359614

**Authors:** Maria A. Gutierrez, Caroline K. Page, Stephen M. Tompkins, Pejman Rohani

## Abstract

The coexistence of competing pathogen strains is shaped by cross-immunity, the cross-protection that infection with one strain confers against another. Although cross-immunity is often asymmetric between strains, this asymmetry is often neglected in the literature on multi-strain coexistence. The effect on coexistence-exclusion outcomes of waning immunity—which is particularly relevant for antigenically evolving pathogens—is also poorly understood. To understand how these factors affect strain coexistence, here we analyze a status-based two-strain SIRS model with asymmetric cross-immunity and strain-specific rates for transmission, recovery, and waning of immunity. We derive explicit invasion thresholds that also determine the feasibility and local stability of a unique coexistence equilibrium. Thus, these thresholds allow us to characterize the region of stable strain coexistence, as a function of the cross-immunities and rates of waning immunity. We also obtain closed-form expressions for the strain prevalences at the coexistence equilibrium, showing that the total prevalence may vary non-monotonically as the basic reproduction number of one strain increases. Finally, we show that a transient reduction in transmission can move a coexisting strain pair across an invasion boundary, driving the weaker strain extinct. Applying this result to influenza B, our analysis offers a parsimonious explanation for the disappearance of the Yamagata lineage during the COVID-19 pandemic.

## 1 Introduction

The interaction of distinct pathogen strains often shapes the ecology, evolution, and epidemiology of many infectious diseases [4, 15, 21], such as human influenza [27] and dengue [1, 8, 34]. When strains interact through cross-immunity (whereby infection with one strain protects against another), the strains compete for susceptible hosts [21]. The outcome of this competition—exclusion of all but one strain, or stable coexistence of several—depends critically on the balance between transmission advantage and immune-mediated exclusion [30, 35]. The competitive exclusion principle establishes that under complete cross-immunity, the strain with the largest basic reproduction number (the average number of infections caused by a single infected host in an otherwise fully-susceptible population [21]) excludes all others [5]. However, partial (rather than complete) cross-immunity can lead to coexistence [35].

Much of the classical literature on strain coexistence [2, 7, 11, 22] implicitly assumes symmetric cross-immunity: the protection that infection with one strain confers against another strain is assumed equal to the protection conferred in the reverse direction. Relaxing this assumption allows for *asymmetric* cross-immunity: differences in the level of protection elicited by one strain to another depending on the infecting strain. There is evidence of asymmetric cross-immunity between *Bordetella pertussis* and *B. parapertussis* [36] and between SARS-CoV-2 variants [28]. Cross-immunity is also asymmetric between B/Victoria and B/Yamagata (the two lineages of the influenza B virus lineages that co-circulated globally from the early 2000s until 2020 [26, 33]): in ferrets, Victoria elicits a stronger cross-reactive response against Yamagata than the reverse [25]. Since the COVID-19 lockdowns started in March 2020 [17], the Yamagata lineage has not been isolated and is considered extinct [6]. In contrast, Victoria reemerged in late 2021 as restrictions were relaxed [6]. These divergent post-pandemic outcomes between the two influenza B lineages may be explained in part by asymmetric cross-immunity [25], thus highlighting the need to explore its effects on strain competition.

A second simplifying assumption common in analyses of strain coexistence [2, 5, 7, 9, 11, 22, 35] is lifelong immunity following infection. However, immunity can wane over months to years: waning of immunity occurs in many pathogens, including SARS-CoV-2 [13, 29] and influenza [37]. Moreover, host protection to these pathogens decrease over time due to their antigenic evolution at the population level [3, 20, 23, 27]. Since different pathogen strains or lineages may be under different selection pressures [33], the rate of waning may also differ between them, potentially affecting coexisting outcomes. A strain that induces shorter-lived immune protection replenishes faster its pool of susceptible hosts, partially compensating for any competitive disadvantage in transmissibility or cross-immunity. A two-strain model incorporating both asymmetric immunity periods and partial cross-immunity recently showed that disparities in temporary immunity periods alone can confer a significant competitive advantage even when strains share similar basic reproduction numbers [19]. The analysis of [19], however, assumed that only one strain confers (partial) cross-immunity: the other strain does not protect at all against the former. Thus, strain coexistence in the general scenario of asymmetric partial cross-immunity and waning immunity has not yet been studied.

In this paper, we characterize how waning immunity and asymmetric cross-immunity jointly shape the ecological competition between two pathogen strains. We analyze a status-based [12] two-strain SIRS model with asymmetric cross-immunity and strain-specific rates of transmission, recovery, and waning of immunity. We derive the invasion thresholds that determine the feasibility and stability of the coexistence equilibrium. We show how the boundaries of the coexistence region depend on the cross-immunity asymmetry and the relative durations of immunity. We also derive the combined disease prevalence at the coexistence equilibrium, and show that it can vary non-monotonically with the basic reproduction number of a strain. We then apply the model to influenza B, showing that a transient reduction in transmission (of the kind imposed during the COVID-19 pandemic) can move a previously coexisting pair of strains across a coexistence boundary, driving one lineage to extinction while the other persists.

## 2 Methods

### 2.1 Epidemic model

We use a status-based [12] compartmental epidemiological model for two pathogen strains: Y and V. Immunity is indexed by the strain against which it protects rather than by the strain that elicited it. Each strain follows *SIRS* dynamics, with *S*_*V*_ and *I*_*V*_ denoting the fraction of the population susceptible to and infectious with strain V, and similarly for *S*_*Y*_ and *I*_*Y*_ . As in [11], we assume that strain cross-immunity is all-or-none [14]: the protection against Y provided by infection with V is either complete immunity (with probability *σ*_*V Y*_ ) or inexistent (with probability 1 −*σ*_*V Y*_ ). Similarly, *σ*_*Y V*_ denotes the protection from Y to V (we allow for potentially asymmetric cross-immunity, i.e., *σ*_*V Y*_ ≠ *σ*_*Y V*_ ). We also allow strain-specific rates of disease transmission (*β*_*V*_ and *β*_*Y*_ ), recovery (*γ*_*V*_ and *γ*_*Y*_ ), and waning of immunity (*ω*_*Y*_ and *ω*_*V*_ ). The system of ordinary differential equations is

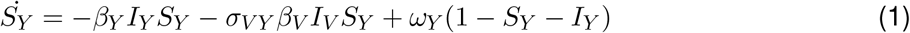

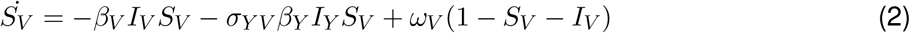

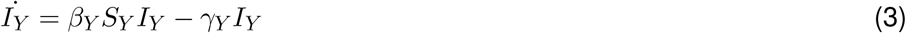

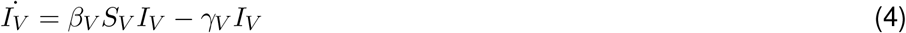

so the basic reproduction numbers [21] for strains V and Y are 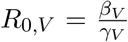 and 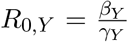. There is one important limitation in the scope of this model. This model is not biologically realistic if the average duration of immunity to one strain (*T*_*V*_ = 1*/ω*_*V*_ or *T*_*Y*_ = 1*/ω*_*Y*_ ) are of comparable magnitude to the respective average infectious period (1*/γ*_*V*_ or 1*/γ*_*Y*_ ). Individuals immune to one strain (say Y) due to infection with the other strain (V) joined the recovered compartment (*R*_*Y*_ = 1 −*S*_*Y*_ −*I*_*Y*_ ) the moment they became infected (with V). Thus, according to the model, while they are infected (in *I*_*V*_ ) their immunity to Y is already waning (since they are in *R*_*Y*_ ), but not their immunity to V (until they recover and join *R*_*V*_ ).

### 2.2 Disease-free equilibrium, single-strain equilibrium, and invasion condition

If *R*_0,*V*_, *R*_0,*Y*_ *<* 1, the disease-free equilibrium (*I*_*V*_ = *I*_*Y*_ = 0) is stable, as in standard epidemiological theory [21]. If *R*_0,*V*_ *>* 1, there exists an endemic equilibrium where only strain V is circulating, analogous to the equilibrium of a single-strain *SIRS* model [21]:

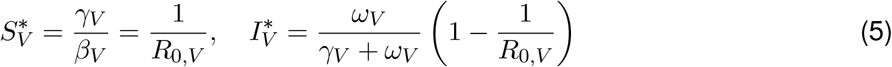

When the V-only equilibrium exists, it is stable if and only if strain Y cannot invade it. The presence of strain V reduces the susceptible pool for Y through cross-immunity. Setting 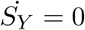 with *I*_*Y*_ = 0 gives the available susceptible pool for Y:

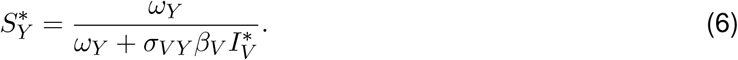

The effective reproduction number of Y at the V-only equilibrium is

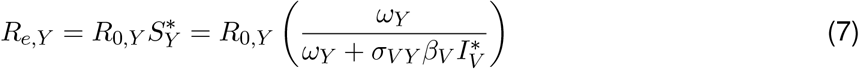

and for strain Y to successfully invade, we need *R*_*e,Y*_ *>* 1, i.e.,

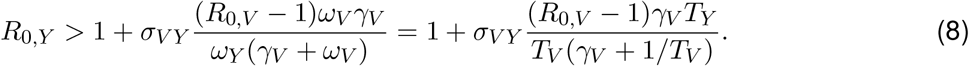

Intuitively, the higher the basic reproduction of V is, the higher the basic reproduction of Y needs to be for it to invade (unless V does not elicit any cross-immunity against Y, i.e., *σ*_*V Y*_ = 0). Similarly, increasing the cross-immunity *σ*_*V Y*_ increases the minimum *R*_0,*Y*_ required for invasion. Increasing the average duration of immunity to V decreases the minimum *R*_0,*Y*_, as this results in fewer V-infections—which indirectly reduce the pool of susceptibles to Y (through cross-immunity *σ*_*V Y*_ *>* 0). In contrast, a longer average duration of immunity to Y requires a higher *R*_0,*Y*_ for Y-invasion, as reinfections occur less often.

By symmetry, if *R*_0,*Y*_ *>* 1 there exists a Y-only endemic equilibrium—given by expressions analogous to (6)—and is invaded by strain V if

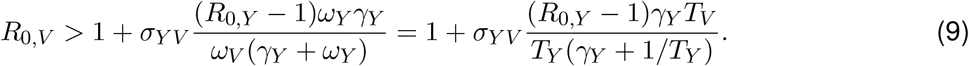

### 2.3 Coexistence equilibrium

### Feasibility

We now search for a coexistence equilibrium, *E*_*co*_ = (*Ŝ*_*V*_, *Î*_*V*_, *Ŝ*_*Y*_, *Î*_*Y*_ ) with *Î*_*V*_, *Î*_*Y*_ *>* 0. From *İ*_*Y*_ = 0 = *İ*_*V*_, we find

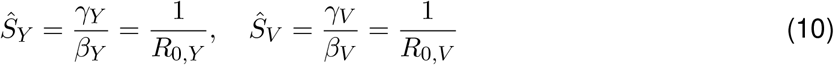

To find *Î*_*Y*_ and *Î*_*V*_, we substitute *Ŝ*_*Y*_ and *Ŝ*_*V*_ into the equations for 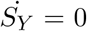 and 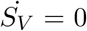. Rearranging the terms yields a 2 × 2 linear system:

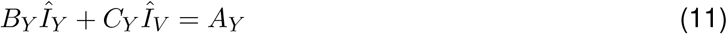

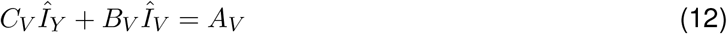

where

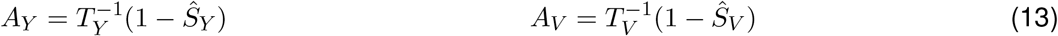

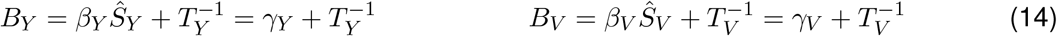

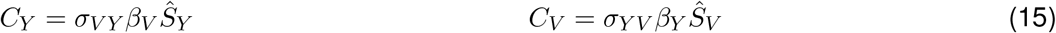

Since *B*_*Y*_ *B*_*V*_ *> C*_*Y*_ *C*_*V*_ for all *σ*_*V Y*_ *σ*_*Y V*_ *<* 1 (cross-immunity is imperfect in at least one direction),

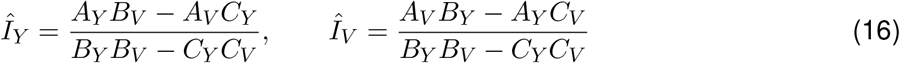

provided that *A*_*Y*_ *B*_*V*_ *> A*_*V*_ *C*_*Y*_ and *A*_*V*_ *B*_*Y*_ *> A*_*Y*_ *C*_*V*_ . The expression in (16) for *Î*_*Y*_ is positive if and only if the condition (8) for the invasion of Y holds. Similarly, *Î*_*V*_ is positive if and only if V can invade the Y-only endemic equilibrium. Thus, the coexistence equilibrium is biologically feasible (*Î*_*V*_, *Î*_*Y*_ *>* 0) if and only if both (8) and (9) hold. Moreover, this derivation also shows that the coexistence equilibrium is unique.

### Total prevalence

The expressions (16) also show that the total strain prevalence, *Î*_*V*_ + *Î*_*Y*_, as a function of *R*_0,*Y*_, has a local maximum at

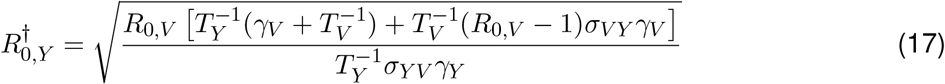

provided that 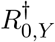(as above) and *R*_0,*V*_ (fixed) lead to strain coexistence, i.e.,

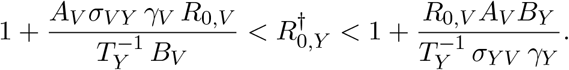

In other words, when 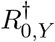 falls within the coexistence window, the total prevalence is a non-monotonic function of *R*_0,*Y*_ . Whether 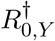 is a local or a global maximum depends on *R*_0,*V*_ (Figure 1). For small 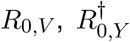 lies above the coexistence window and the feasible total prevalence increases monotonically, with no interior maximum (left panel of Figure 1). For intermediate 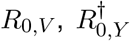 is a local maximum, but larger values of *R*_0,*Y*_ eventually generate an even greater total prevalence (middle panel). For large *R*_0,*V*_, the total prevalence at 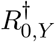 is the global maximum (right panel).

**Figure 1.**
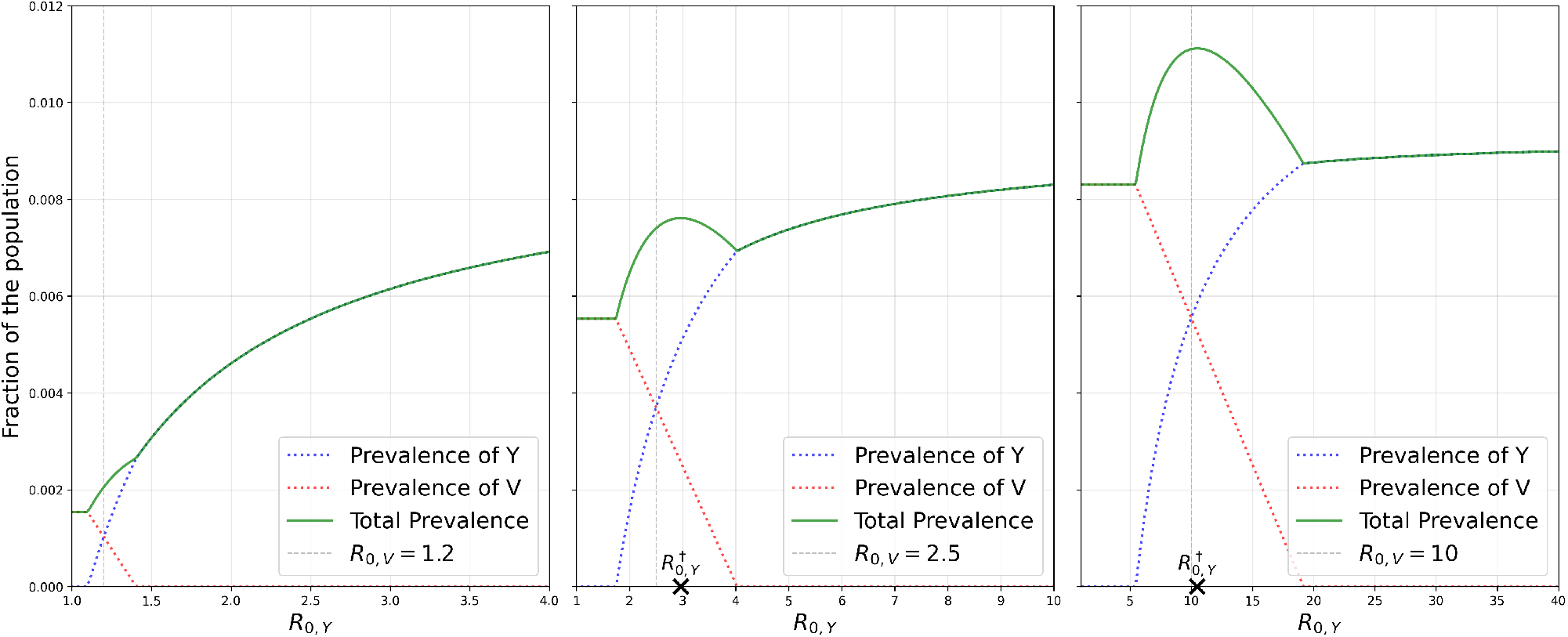
Total equilibrium prevalence as R_0,Y_ varies, for three values of R_0,V_ (all panels share σ_V Y_ = σ_Y V_ = 0.5, T_V_ = T_Y_ = 1 year, 1/γ_V_ = 1/γ_Y_ = 3.4 days). The local maximum 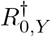is marked when it falls within the coexistence window. Left 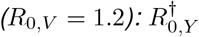lies above the coexistence window, so the total prevalence increases monotonically. Middle 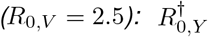is a local maximum, but larger R_0,Y_ eventually generate an even greater total prevalence, so 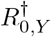is not a global maximum. Right 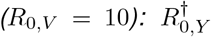 is the global maximum, and the total prevalence there exceeds the single-strain prevalence (which is bounded above by its asymptotic value as R_0,Y_, →∞ 1/(γ_Y_ T_Y_ + 1) ). In all panels, strain Y overtakes V in prevalence once R_0,Y_ > R_0,V_, since all other parameters are shared between the strains.

### Local stability

We first evaluate the Jacobian matrix [24] at the coexistence equilibrium *E*_*co*_ = (*Ŝ*_*V*_, *Î*_*V*_, *Ŝ*_*Y*_, *Î*_*Y*_ ):

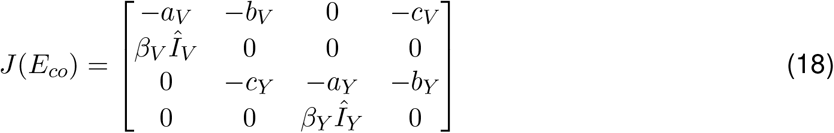

where, for (*i, j*) = (*V, Y* ), (*Y, V* ),

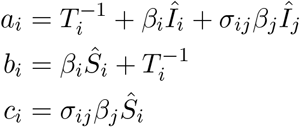

are all strictly positive constants. Expanding the determinant det(*J* (*E*_*co*_) −*λI*) yields a fourth-degree characteristic polynomial:

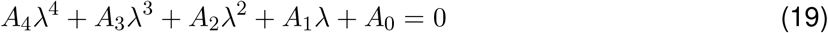

Where

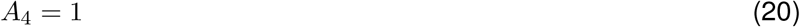

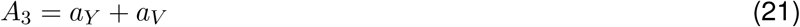

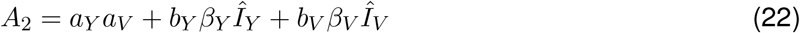

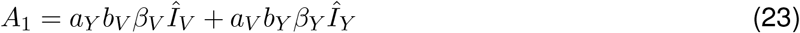

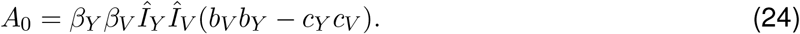

According to the Routh-Hurwitz criterion [24], the coexistence equilibrium *E*_*co*_ is locally asymptotically stable if and only if all the following conditions hold:

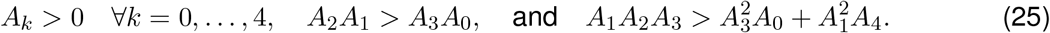

It is straightforward (but tedious) to confirm that all the above are satisfied because *σ*_*V Y*_ *σ*_*Y V*_ *<* 1. Therefore, the coexistence equilibrium is locally asymptotically stable.

### 2.4 Phase diagrams

The above stability results show that, for a given set of parameters, only one equilibrium point is stable. Thus, parameter space is split into four regions, corresponding to (i) the disease-free equilibrium if *R*_0,*V*_, *R*_0,*Y*_ *<* 1, (ii) the V-only endemic equilibrium if *R*_0,*V*_ *>* 1 but (8) does not hold, (iii) the Y-only endemic equilibrium if *R*_0,*Y*_ *>* 1 but (9) does not hold, and (iv) the coexistence equilibrium if both (8) and (9) hold. These regions yield the phase diagrams of Figure 2.

**Figure 2.**
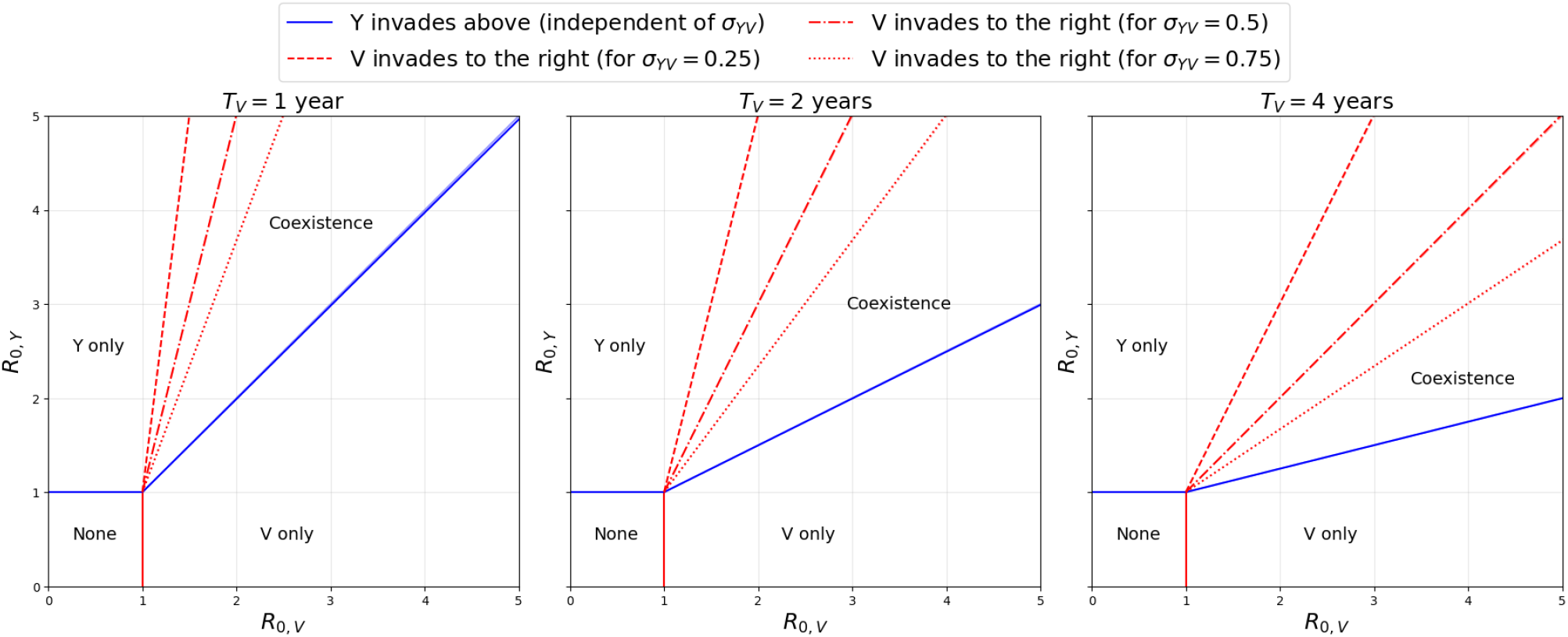
Phase diagrams determining which strain persists, depending on the basic reproduction numbers R_0,V_ and R_0,Y_ . Fixed σ_V Y_ = 0.5, T_Y_ = 2 years, 1/γ_V_ = 1/γ_Y_ = 3.4 days.

As mentioned earlier, the model is biologically feasible provided that *T*_*V*_ ≫ 1*/γ*_*V*_ and *T*_*Y*_ ≫ 1*/γ*_*Y*_ . These limits give approximations for (8) and (9):

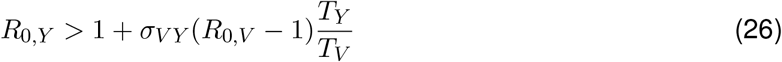

and

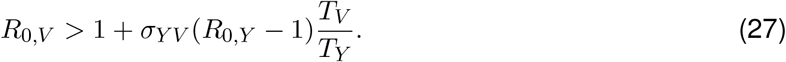

The boundaries generated by these approximated conditions are also included in Figure 2, with a lower transparency than the exact boundaries, but due to their large overlap the approximated boundaries are barely noticeable (see top right corner of the first plot). In 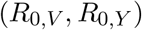-space, the approximated boundary (26) for Y-invasion is a straight line (blue) with slope 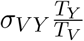 (for *R*_0,*V*_ *>* 1). Similarly, the boundary for V-invasion (27) is a line (red) with slope 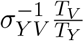 . Between these two lines, both strains coexist.

Increasing any of the cross-immunity parameters shrinks the region of coexistence, as intuitively expected. For example, increasing the cross-immunity *σ*_*Y V*_ reduces the slope of the line determining the threshold for Y invasion: the red lines in each of the subplots of Figure 2 tilt to the right. In contrast, the threshold for Y invasion (blue line) remains constant as the cross-immunity *σ*_*Y V*_ increases. Increasing the duration of immunity to V rotates both lines in the same direction: this is seen across the subplots of Figure 2. The duration of immunity to V affects its potential for invasion because with slower waning, hosts immune to V due to cross-immunity take longer to return to the susceptible pool. Hence, longer duration of immunity requires a higher *R*_0,*V*_ for invasion (red line). With slower waning, the prevalence of V is lower, generating less cross-immunity to Y. Therefore, the minimum *R*_0,*Y*_ for Y invasion is lower (blue line).

## 3 Application to the extinction of influenza B/Yamagata

We now apply the model to the apparent extinction of the Yamagata lineage during the COVID-19 pandemic. We let strain Y represent Yamagata and strain V represent Victoria, the lineage that resumed normal circulation once pandemic restrictions were relaxed [6]. The two lineages had co-circulated globally for roughly two decades prior to 2020 [33]. In our model, we equate the pre-pandemic coexistence of Victoria and Yamagata to the stable coexistence equilibrium *E*_*co*_.

We model the COVID-19 lockdowns as a temporary reduction *c*_0_ in contacts, so that during the period of reduced transmission the basic reproduction numbers become *R*_0,*V*_ (1 − *c*_0_) and *R*_0,*Y*_ (1 − *c*_0_). Because the inequalities (8) and (9) are not invariant under rescaling of the reproduction numbers, a reduction that preserves the ratio *R*_0,*Y*_ */R*_0,*V*_ can move the system from one stable equilibrium to another. In particular, a strain pair that coexists at pre-pandemic transmission levels can be moved across the invasion threshold for the weaker competitor, driving it to extinction.

To apply this analysis to Victoria and Yamagata, we assume that Victoria induces a stronger cross-immunity to Yamagata than vice versa (*σ*_*V Y*_ *> σ*_*Y V*_ ), as suggested by challenge experiments in ferrets [25]. In particular, for Figure 3, we use values for the cross-immunities based on the data reported in [25], with ferrets infected with either Victoria or Yamagata, six months after being infected with the other lineage. Table 1 reports viral titers for the four animal subjects on the first day of infection. To calculate the cross-immunity from Victoria to Yamagata, we divide the average Yamagata infectivity (assumed proportional to the logarithm of the viral load [18] on the first day of infection) across the four ferrets that had previously been infected with Victoria by the average infectivity (using the same logarithmic formulation) of four naïve ferrets challenged with Yamagata. One minus this ratio gives the cross-immunity *σ*_*V Y*_ = 0.46, and an analogous calculation yields *σ*_*Y V*_ = 0.15. This calculation implicitly assumes that cross-immunity reduces infectivity, not susceptibility, which would be consistent with our model assumption. That said, in multi-strain systems, it is usually unimportant for the dynamical output which form of cross-immunity is used [9].

**Table 1.** Viral loads value (plaque-forming units per mL) from the experiments reported in [25], used to calculate the cross-immunity parameters σ_V Y_ and σ_Y V_ between the influenza B/Victoria and B/Yamagata lineages.

| Infecting strain | Immune state | Viral titers (PFU/mL) |  |  |  |
| --- | --- | --- | --- | --- | --- |
| Yamagata | Victoria infection six months earlier | 400 | 220 | 400 | 40 |
|  | Naïve | 25000 | 20000 | 10000 | 16000 |
| Victoria | Yamagata infection six months earlier | 14000 | 500 | 11000 | 3000 |
|  | Naïve | 6000 | 11000 | 6000 | 170000 |

**Figure 3.**
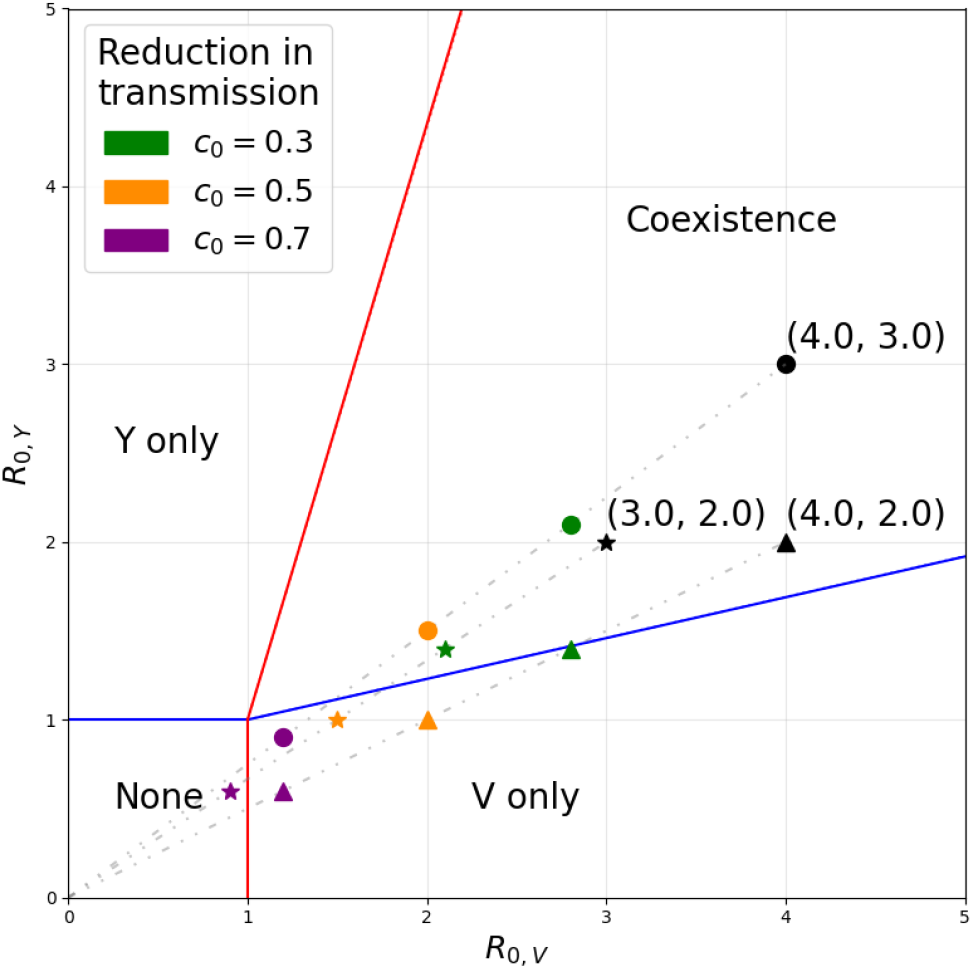
Phase diagram including the effect of a reduction c_0_ in transmission as a result of the COVID-19 lockdown. Fixed parameters T_V_ = 4 years, T_Y_ = 2 years, σ_V Y_ = 0.46, σ_Y V_ = 0.15, 1/γ_V_ = 1/γ_Y_ = 3.4 days. Three pairs of values for (R_0,V_, R_0,Y_ ) are considered: (4, 3), denoted with circles; (3, 2), denoted with stars; and (4, 2), denoted with triangles.

We also assume that Yamagata is subject to faster antigenic evolution than Victoria, as indicated by phylogenetic analyses [33]. This, in our model, translates to a shorter average duration of immunity to Yamagata than Victoria (*T*_*Y*_ *< T*_*V*_ ), also in agreement with the findings of [31]. Finally, we assume that Victoria has a higher basic reproduction number (*R*_0,*V*_ *> R*_0,*Y*_ ). This assumption is consistent with analyses which estimate a lower *effective* reproduction number for Yamagata than Victoria [32]. We consider values of these reproduction numbers such that, before COVID-19, (8) and (9) predict the stable coexistence of both lineages. Figure 3 shows that the strain balance can shift from coexistence to the Victoria-only endemic equilibrium (resulting in the extinction of Yamagata), for intermediate values of the reduction in transmission *c*_0_. A strong lockdown (high *c*_0_) leads to the disease-free equilibrium (both lineages extinct), while a mild lockdown (low *c*_0_) maintains coexistence. The exact threshold value of the strength of the lockdown *c*_0_ required for the extinction of Yamagata depends on the other parameters.

## 4 Discussion

We have analyzed a status-based two-strain *SIRS* model with asymmetric partial cross-immunity and strain-specific waning immunity. We have derived analytical expressions for the invasion thresholds that determine the feasibility and stability the coexistence equilibrium, thus generating phase diagrams with the possible coexistence-exclusion outcomes. We have also obtained analytical expressions for the prevalence of each strain at the coexistence equilibrium. Applying the model to influenza B, we have shown that a transient reduction in transmission can displace a coexisting pair of strains across a coexistence boundary and drive the weaker competitor to extinction, offering a parsimonious dynamical explanation for the disappearance of one of the two co-circulating influenza B lineages during the COVID-19 pandemic.

For the analysis in the main text, we have assumed all-or-none cross-immunity [11]. The all-or-none assumption allows for derivation of closed-form expressions for the prevalences at the coexistence equilibrium and leads to a tractable analysis of local stability, in contrast with the largely numerical treatment when cross-immunity is assumed to be “leaky” (e.g. [19]). Moreover, the status-based formulation [12] also leads to analytical thresholds for strain invasion (minimum values of the basic reproduction numbers *R*_0,*V*_ and *R*_0,*Y*_ ), with both all-or-none immunity (main text) and leaky immunity [11] (Section 5.1). For the former, we have found that the boundary of the coexistence region in (*R*_0,*V*_, *R*_0,*Y*_ )-space consists of two straight lines (representing the invasion thresholds), whose slopes are set by the cross-immunities and the ratio of the average durations of immunity. This tractability not only makes clear the dependence on each parameter, it also underscores the importance of considering strain-specific parameters for the duration of immunity and the (asymmetric and partial) cross-immunity. Both of waning immunity and asymmetric cross-immunity have been traditionally neglected in the multi-strain literature. Including them in our model, allows us to recapitulate the extinction of the B/Yamagata influenza lineage during the COVID-19 pandemic. In particular, a temporary reduction in transmission moves the systems from a two-strain coexistence equilibrium (pre-pandemic) to a single-lineage equilibrium (corresponding to the persistence of the B/Victoria lineage).

Two other results stand out from our analysis. First, the type of cross-immunity qualitatively changes how a transmission advantage maps onto competitive outcome. Under all-or-none cross-immunity, the invasion thresholds (8)–(9) are, in the biologically relevant limit *T* ≫ 1*/γ*, straight lines through the point (1, 1) in (*R*_0,*V*_, *R*_0,*Y*_ )-space (Figure 2). Coexistence is then confined to a wedge between these lines, and a strain with a sufficiently large reproduction number can always exclude its competitor. In contrast, under leaky cross-immunity the invasion boundaries instead bend towards vertical and horizontal asymptotes (Figure 4). Therefore, under partial leaky cross-immunity, strains may coexist even when one strain is far more transmissible than the other. This ‘revision’ of the competitive exclusion principle was also found in a similar model with leaky model [10]). The contrasting results for all-or-none immunity, however, suggests that under more general mechanisms of strain cross-immunity [14], the competitive exclusion principle may be recovered (a large enough transmission advantage guarantees exclusion).

**Figure 4.**
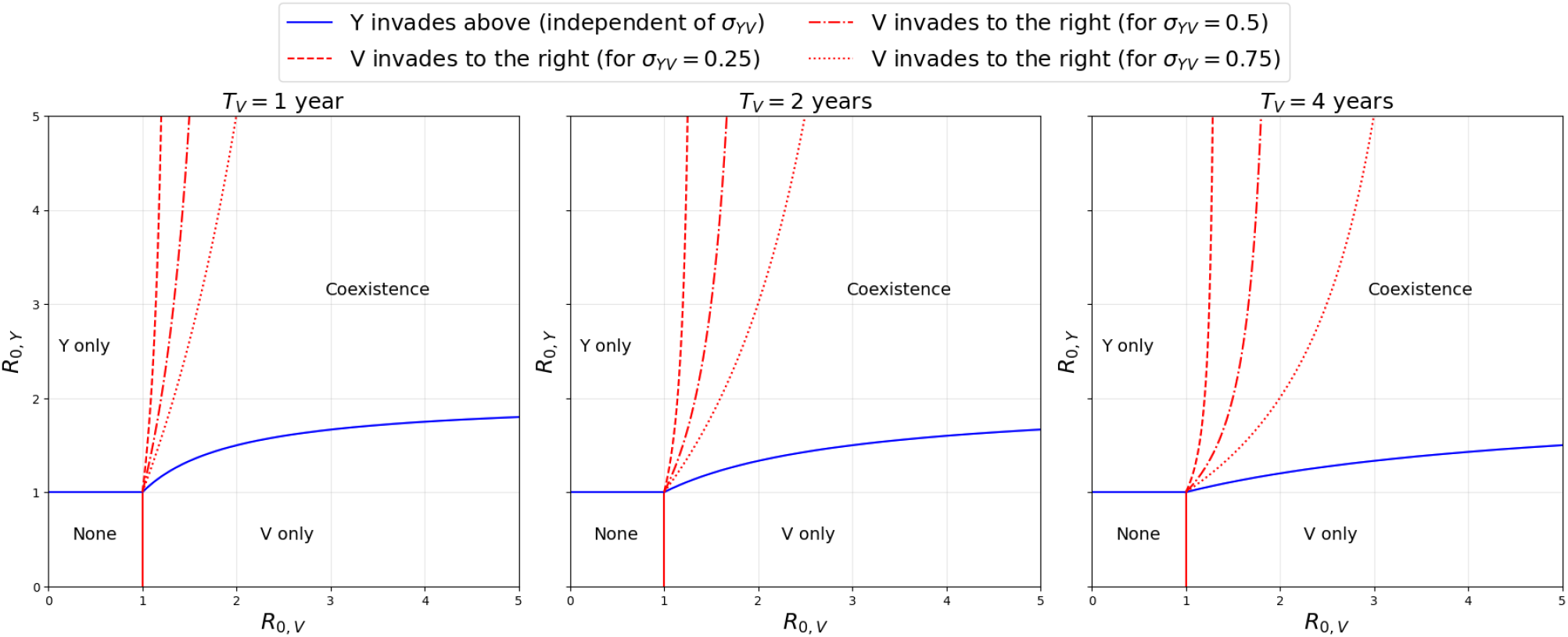
Coexistence diagrams for leaky immunity, instead of all-or-none. While the parameters are as in Figure 2, leaky immunity leads to vertical and horizontal asymptotes.

**Figure 5.**
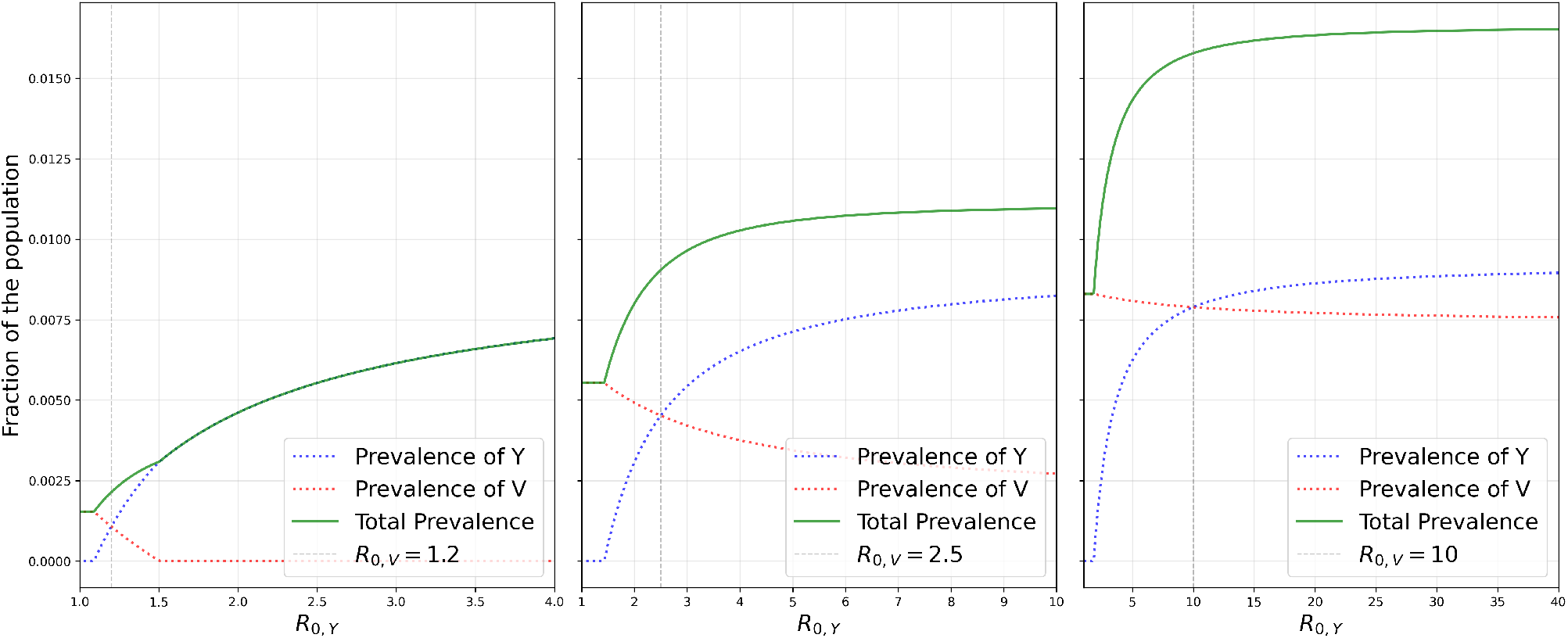
Total equilibrium prevalence as R_0,Y_ varies, under leaky cross-immunity, for the same three parameter sets as Figure 1. Unlike the all-or-none case (Figure 1), the total prevalence always increases monotonically with R_0,Y_ .

Second, under all-or-none cross-immunity the combined prevalence of the two strains may vary non-monotonically with the basic reproduction number of one strain. In particular, as that reproduction number (say *R*_0,*Y*_ ) increases, the total prevalence at the coexistence equilibrium reaches a local maximum for intermediate or high values of the other strain basic reproduction number (*R*_0,*V*_ ). For intermediate values of *R*_0,*V*_, as *R*_0,*Y*_ → ∞, the single-strain prevalence eventually surpasses the prevalence at the local maximum. With high values of *R*_0,*V*_, the local maximum at the coexistence equilibrium is a global maximum of the total prevalence. For low values of *R*_0,*V*_, the total prevalence increases monotonically with *R*_0,*Y*_ . Under leaky cross-immunity, however, the total prevalence always increases monotonically with *R*_0,*Y*_, regardless of *R*_0,*V*_ . These qualitatively different outcomes depending on the mechanism of immunity are reminiscent of the different stability results found for other pairs of models with leaky and all-or-none lifelong immunity [7, 16].

When including waning immunity, there is a subtle caveat to the status-based formulation. It implictly assumes that a host immune to Y through infection with V begins to lose that protection immediately, even while still infectious with V. Consequently, when the average duration of immunity is not much longer than the infectious period, the full invasion conditions (8) and (9) permit the coexistence of a strictly weaker strain—identical to its competitor in every parameter except for slightly lower transmissibility—even under complete cross-immunity. This contradiction of the competitive exclusion principle [5] disappears in the biologically-relevant regime in which, for each strain, the average duration of immunity is much longer than the average duration of infection.

In the main text, we have also neglected vital (birth and death dynamics), as we expect the turnover of the host population to be relevant over a longer timescale than the durations of immunity considered here. Nevertheless, in the Appendix (Section 5.2) we show that adding demography at constant population size, as in [11], keeps the linear structure in the invasion thresholds and preserves the closed-form coexistence equilibrium. Another natural extension to our model would be to include vaccination, which can reshape the coexistence boundary [35]. We have neglected vaccination to focus on the specific effects of waning immunity and asymmetric cross-immunity, but it would be straightforward to include vaccination against one or both strains in our model formulation. A more challenging extension of this work would be to consider more than two strains, which, at least without waning immunity, can lead to oscillatory coexistence (periodic orbits) [2, 7].

## Data Availability

All data produced in the present work are contained in the manuscript

## 5 Appendix

### 5.1 Leaky immunity

In the main text cross-immunity is all-or-none [11]. Here we show that the alternative assumption, leaky cross-immunity, yields the same qualitative coexistence structure, but with invasion thresholds that are no longer linear in the reproduction numbers.

For the invasion analysis, we consider the force of infection 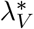 at the V-only equilibrium (6), and classify a host by its status with respect to Y. Thus, the model here is still status-based ([12]). We include compartments for fully susceptible hosts (*S*), partially immune through prior V-infection (*P*, with susceptibility reduced by a factor 1 −*σ*, where *σ* ≡ *σ*_*V Y*_ ), or infected from either compartment (*I*_*S*_, *I*_*P*_). Moreover, we assume that a fraction 1 −*S* −*P* −*I*_*S*_ −*I*_*P*_ of the population are fully immune to strain 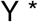 (following recovery), but this infection-induced immunity still wanes at the same rate *ω*_*Y*_ of the cross-immunity. Hence, the ODEs for the transmission of strain Y are

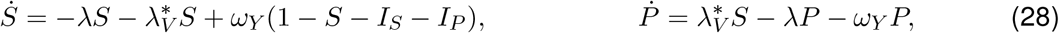

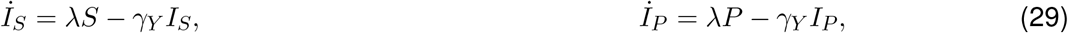

with *λ* = *β*_*Y*_ *I*_*S*_ + (1 −*σ*)*I*_*P*_ . When Y is absent (*I*_*S*_ = *I*_*P*_ = 0), the population settles at *S*^∗^ + *P* ^∗^ = 1 with

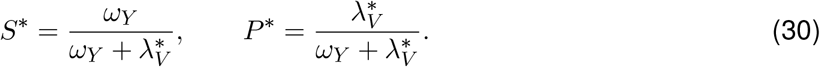

The effective reproduction number of Y at this V-only equilibrium follows from a next-generation-matrix calculation [21]: a host infected from *S* transmits at rate *β*_*Y*_ and one infected from *P* at rate *β*_*Y*_ (1 −*σ*), and both recover at rate *γ*_*Y*_, so

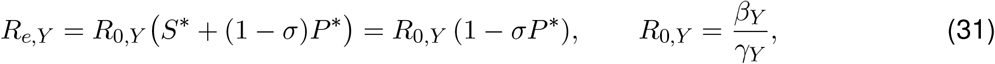

using *S*^∗^ + *P* ^∗^ = 1. Strain Y invades when *R*_*e,Y*_ *>* 1, i.e. *σP* ^∗^ *<* 1 − 1*/R*_0,*Y*_ . Substituting 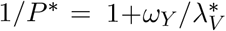 together with the single-strain force of infection 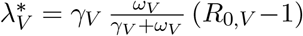 gives the invasion condition

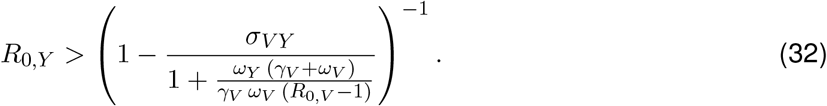

Under the approximation of much shorter average durations of infection than durations of immunity— the same limit that yields the all-or-none condition (26)—the condition becomes

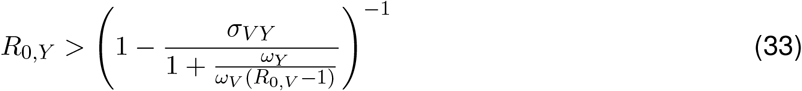

the leaky analogue of (26). Unlike that linear boundary, it asymptotes to 1*/*(1 −*σ*_*V Y*_ ) in the limit *R*_0,_ ≫ _*V*_ 1, so that (for imperfect cross-immunity) coexistence is possible even when one strain is much more transmissible than the other, as in [10].

### 5.2 Birth-and-death dynamics

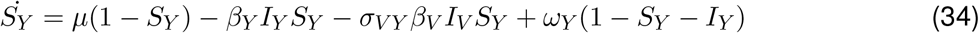

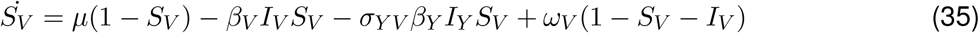

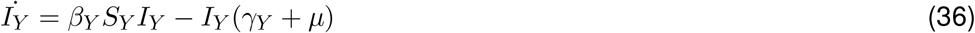

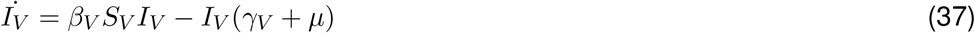

The basic reproduction numbers are now

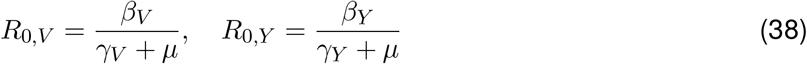

The V-only equilibrium is

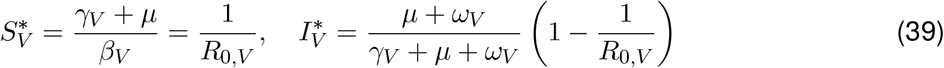

The V-only equilibrium is stable if and only if strain Y cannot invade. The presence of strain V reduces the susceptible pool for Y through cross-immunity. Setting 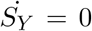 with *I*_*Y*_ = 0 gives the available susceptible pool for Y:

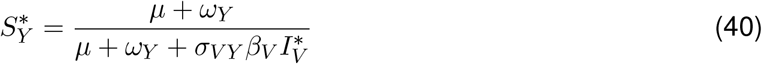

The effective reproduction number of Y at the V-only equilibrium is

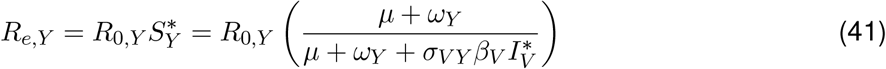

As before, for strain Y to successfully invade, we need *R*_*e,Y*_ *>* 1. Therefore, the condition (8) becomes

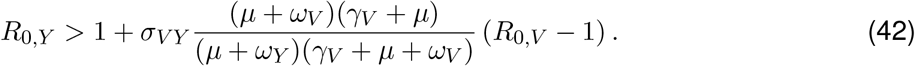

which remains linear on the *R*_0_s. If immunity does not wane (*ω*_*V*_ = 0 = *ω*_*Y*_ ), the condition (42) becomes

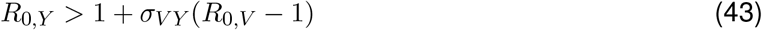

which is equivalent to the invasion condition (4) of [35] (under the change of variables *α*_2_ → *σ*_*V Y*_, *R*_0,2_ → *R*_0,*Y*_, *R*_0,1_ → *R*_0,*V*_ ). By symmetry from (42), the condition (9) for V to invade the Y-only equilibrium becomes

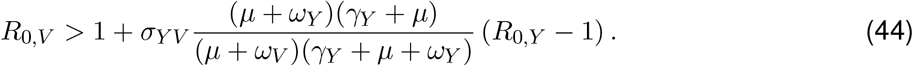

These invasion conditions (42) and (44) remain linear on the *R*_0_*s* and the cross-immunities.

The coexistence equilibrium still has *Ŝ*_*Y*_ = 1*/R*_0,*Y*_, *Ŝ*_*V*_ = 1*/R*_0,*V*_ and prevalences *Î*_*V*_, *Î*_*Y*_ given by (16), where now

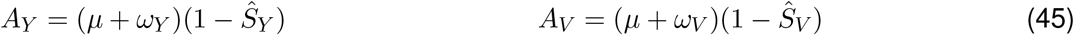

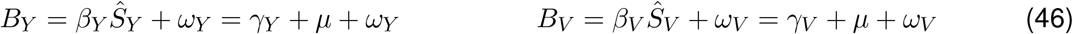

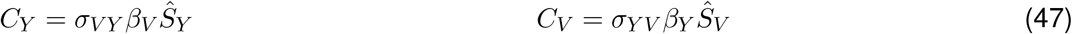

Thus, the coexistence equilibrium is biologically feasible if and only if the invasion conditions (42) and both hold. The local stability analysis is as in Section 2.3.

## Acknowledgements

We thank the Rohani Lab for comments. This project has been funded with Federal funds from the National Institute of Allergy and Infectious Diseases, National Institutes of Health, Department of Health and Human Services, under Contract No. 75N93021C00018 (NIAID Centers of Excellence for Influenza Research and Response, CEIRR).

